# Common variants in Alzheimer’s disease: Novel association of six genetic variants with AD and risk stratification by polygenic risk scores

**DOI:** 10.1101/19012021

**Authors:** Itziar de Rojas, Sonia Moreno-Grau, Niccolò Tesi, Benjamin Grenier-Boley, Victor Andrade, Iris Jansen, Nancy L. Pedersen, Najada Stringa, Anna Zettergren, Isabel Hernández, Laura Montrreal, Carmen Antúnez, Anna Antonell, Rick M. Tankard, Joshua C. Bis, Rebecca Sims, Céline Bellenguez, Inés Quintela, Antonio González-Perez, Miguel Calero, Emilio Franco, Juan Macías, Rafael Blesa, Manuel Menéndez-González, Ana Frank-García, Jose Luís Royo, Fermín Moreno, Raquel Huerto, Miquel Baquero, Mónica Diez-Fairen, Carmen Lage, Sebastian Garcia-Madrona, Pablo García, Emilio Alarcón-Martín, Sergi Valero, Oscar Sotolongo-Grau, EADB, GR@ACE, DEGESCO, IGAP (ADGC, CHARGE, EADI, GERAD) and PGC-ALZ Consortia, Guillermo Garcia-Ribas, Pascual Sánchez-Juan, Pau Pastor, Jordi Pérez-Tur, Gerard Piñol-Ripoll, Adolfo Lopez de Munain, Jose María García-Alberca, María J. Bullido, Victoria Álvarez, Alberto Lleó, Luis M. Real, Pablo Mir, Miguel Medina, Philip Scheltens, Henne Holstege, Marta Marquié, María Eugenia Sáez, Ángel Carracedo, Philippe Amouyel, Julie Williams, Sudha Seshadri, Cornelia M. van Duijn, Karen A. Mather, Raquel Sánchez-Valle, Manuel Serrano-Ríos, Adelina Orellana, Lluís Tárraga, Kaj Blennow, Martijn Huisman, Ole A. Andreassen, Danielle Posthuma, Jordi Clarimón, Mercè Boada, Wiesje M. van der Flier, Alfredo Ramirez, Jean-Charles Lambert, Sven J. van der Lee, Agustín Ruiz

**Author notes:** **Corresponding authors:** Agustín Ruiz M.D. Ph.D., Address: Research Center. Fundació ACE. Institut Català de Neurociències Aplicades., C/ Marquès de Sentmenat, 57. 08029 Barcelona, Spain, Email id, Sven J. van der Lee M.D. Ph.D., Alzheimer Center Amsterdam, Department of Neurology, Amsterdam Neuroscience, Vrije Universiteit Amsterdam, Amsterdam UMC, Amsterdam, The Netherlands, Address: De Boelelaan 1117, 1081 HV, Amsterdam., Email id. These authors contributed equally to this work (Itziar de Rojas, Sonia Moreno-Grau, Niccolò Tesi, Benjamin Grenier-Boley, Victor Andrade, Iris Jansen). These authors jointly supervised this work (Jordi Clarimón, Mercè Boada, Wiesje M. van der Flier, Alfredo Ramirez, Jean-Charles Lambert, Sven J. van der Lee, Agustín Ruiz). **Conflict of Interest:** None.

## Abstract

**BACKGROUND:** Disentangling the genetic constellation underlying Alzheimer’s disease (AD) is important. Doing so allows us to identify biological pathways underlying AD, point towards novel drug targets and use the variants for individualised risk predictions in disease modifying or prevention trials. In the present work we report on the largest genome-wide association study (GWAS) for AD risk to date and show the combined utility of proven AD loci for precision medicine using polygenic risk scores (PRS).

**METHODS:** Three sets of summary statistics were included in our meta-GWAS of AD: an Spanish case-control study (GR@ACE/DEGESCO study, n = 12,386), the case-control study of International Genomics of Alzheimer project (IGAP, n = 82,771) and the UK Biobank (UKB) AD-by-proxy case-control study (n=314,278). Using these resources, we performed a fixed-effects inverse-variance-weighted meta-analysis. Detected loci were confirmed in a replication study of 19,089 AD cases and 39,101 controls from 16 European-ancestry cohorts not previously used. We constructed a weighted PRS based on the 39 AD variants. PRS were generated by multiplying the genotype dosage of each risk allele for each variant by its respective weight, and then summing across all variants. We first validated it for AD in independent data (assessing effects of sub-threshold signal, diagnostic certainty, age at onset and sex) and tested its effect on risk (odds for disease) and age at onset in the GR@ACE/DEGESCO study.

**FINDINGS:** Using our meta-GWAS approach and follow-up analysis, we identified novel genome-wide significant associations of six genetic variants with AD risk (rs72835061*-CHRNE*, rs2154481*-APP*, rs876461*-PRKD3/NDUFAF7*, rs3935877*-PLCG2* and two missense variants: rs34173062/rs34674752 in *SHARPIN* gene) and confirmed a stop codon mutation in the *IL34* gene increasing the risk of AD (*IL34-Tyr213Ter*), and two other variants in *PLCG2* and *HS3ST1* regions. This brings the total number of genetic variants associated with AD to 39 (excluding *APOE*). The PRS based on these variants was associated with AD in an independent clinical AD-case control dataset (OR=1.30, per 1-SD increase in the PRS, 95%CI 1.18-1.44, *p =* 1.1×10^−7^), a similar effect to that in the GR@ACE/DEGESCO (OR=1.27, 95%CI 1.23-1.32, *p =* 7.4×10^−39^). We then explored the combined effects of these 39 variants in a PRS for AD risk and age-at-onset stratification in GR@ACE/DEGESCO. Excluding *APOE*, we observed a gradual risk increase over the 2% tiles; when comparing the extremes, those with the 2% highest risk had a 2.98-fold (95% CI 2.12–4.18, *p* = 3.2×10^−10^) increased risk compared to those with the 2% lowest risk (*p* = 5.9×10^−10^). Using the PRS we identified *APOE ε33* carriers with a similar risk as *APOE* ε*4* heterozygotes carriers, as well as *APOE ε4* heterozygote carriers with a similar risk as *APOE* ε*4* homozygote. Considering age at onset; there was a 9-year difference between median onset of AD the lowest risk group and the highest risk group (82 *vs* 73 years; *p* = 1.6×10^−6^); a 4-year median onset difference (81 *vs* 77 years; *p* = 6.9×10^−5^) within *APOE* ε4 heterozygotes and a 5.5-year median onset difference (78.5 *vs* 73 years; *p* = 4.6×10^−5^) within *APOE ε4* carriers.

**INTERPRETATION:** We identified six novel genetic variants associated with AD-risk, among which one common *APP* variant. A PRS of all genetic loci reported to date could be a robust tool to predict the risk and age at onset of AD, beyond *APOE* alone. These properties make PRS instrumental in selecting individuals at risk in order to apply preventative strategies and might have potential use in diagnostic work-up.

## Introduction

Alzheimer’s disease (AD) is the most common neurodegenerative disorder affecting elderly populations worldwide^1^. A small fraction of the occurrence of AD in patients can be explained by rare mutations which cause the autosomal dominant forms of AD (< 1%)^2^. For non-familial cases, the genetic contribution to AD risk is estimated to be between 60–80%^3^ and likely consists of a combination of common and rare alleles, each with low to moderate effects on AD risk, gene–gene and gene–environmental interactions.

Thus far, multiple loci associated with AD have been described next to causal mutations in the *PSEN1, PSEN2* and *APP* genes. The most prominent locus, *APOE*, was detected almost 30 years ago using linkage techniques^4^. *APOE* allele ε4 has a strong effect, conferring a threefold increased risk for AD in heterozygous carriers of the ε4 allele and an 8-to-12-fold risk in the homozygous state^5^. After a long and unsuccessful search for additional AD loci, the development of single nucleotide polymorphism (SNP) arrays permitted the design of comprehensive genome-wide association studies (GWAS). Successive waves of analysis and meta-analysis with increasing sample sizes have been performed to disentangle the genetic background of AD. By combining the information from more than 80,000 participants, the International Genetics of Alzheimer’s project (IGAP) recently released the largest meta-analysis of case-control studies reported to date^6^. In parallel with this case-control analysis, by-proxy AD case-control datasets of Alzheimer’s disease have successfully been used to increase the statistical power of previous AD GWAS^7^. These approaches use the UK Biobank (UKB)^8^, a cohort study of over half a million individuals in the UK in which the history of dementia for the parents is used instead of traditional case-control studies^9^. The by-proxy strategy confirmed the loci identified by IGAP and identified additional candidate loci previously undetected by conventional case-control approaches^9,10^. Overall, in addition to *APOE*, there have to date been identified more than 30 loci have been identified to date that modify the risk of AD^11–16^. These signals, combined with ‘subthreshold’ common variant associations, account for ∼31% of the genetic variance of AD, leaving most of the genetic risk as yet uncharacterised^17^. A meta-study combining all these studies may lead to the identification of associations of genetic loci with AD and might help to confirm loci previously proposed by proxy-AD strategies but requiring additional validations. Larger GWASs are also important to identify the causative variants, pinpoint culprit genes in the AD-associated genomic regions and identify the biological pathways underlying AD.

In addition to the biological insights, disentangling the genetic constellation of common genetic variations underlying AD has clinical relevance. First, genetic associations can point towards novel drug targets. One example is the discovery of the *TREM2* gene, which has led to the development of *TREM2* modulators^17^. These drugs are now being evaluated in trials^18^ in hopes of benefiting patients in the near-distant future. Second, pre-symptomatic AD patients are increasingly included in treatment or prevention trials. To reduce the necessary duration of these costly studies, individuals at high genetic risk of developing the disease are included^19,20^, in clinical trials, such as patients carrying a mutation that causes familial AD and those carrying an *APOE* ε4^18,19^. Other common and rare AD loci are neglected, as the effect or frequency of the individual loci is often small. The combined effect of all these loci, however, could account for a substantial proportion of variation in risk^21^. Indeed, combining the effects of all currently known variants results in a polygenic risk score (PRS) that is associated with conversion of mild cognitive impairment to AD^22,23^ the neuropathological hallmarks of AD, age at onset of disease^24–26^ and lifetime risk of AD^27^. However, the discovery of more variants associated with AD warrant that PRS are updated and again validated. Further it is important to know how diagnostic certainty, age at onset and sex influence the effects of the PRS.

Here we aimed to comprehend and expand the knowledge of the genetic landscape underlying AD. We first performed a meta-GWAS integrating all currently published GWAS case-control data, by-proxy case-control data and the data from the Genome Research at Fundació ACE (GR@ACE) study^28^. We confirm the novel observed associations in a large independent replication study. Then constructed an update of the PRS and test if the effects of the PRS are influenced by diagnostic certainty, sex and age at onset groups. Last, we test if the PRS can be used to identify individuals at highest risk.

## Methods

### Meta-GWAS of AD

This study utilizes the summary statistics from three AD GWAS: the summary statistics calculated from the GR@ACE^28^ case-control study, the International Genomics of Alzheimer project (IGAP)^29^ case-control study and UKB AD-by-proxy case-control study^9^ (Figure 1).

**Figure 1:**
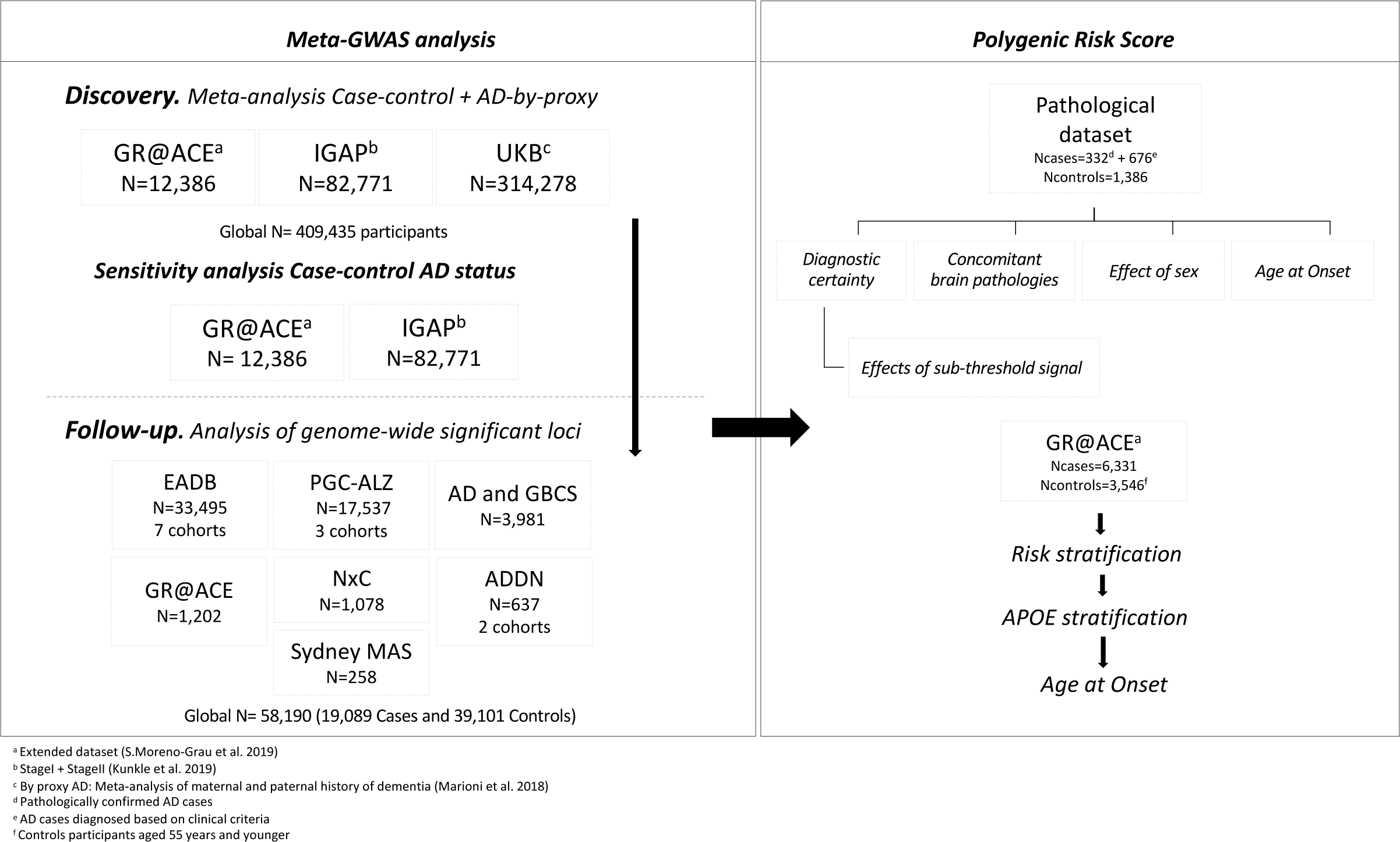
Flow chart of analysis steps. Discovery meta-analysis in GR@ACE, IGAP stage 1+2 and UKBiobank followed by a replication in 16 independent cohorts. The genome-wide significant signals 2 found in meta-GWAS were used to perform a Polygenic Risk Score in a clinical and pathological AD dataset. N = total of individuals within specified data set.

#### GR@ACE

The GR@ACE study^28^ recruited AD patients from Fundació ACE, Institut Català de Neurociències Aplicades (Catalonia, Spain) and control individuals from three centres: Fundació ACE (Barcelona, Spain), Valme University Hospital (Seville, Spain) and the Spanish National DNA Bank Carlos III (University of Salamanca, Spain) (http://www.bancoadn.org). Additional cases and controls were obtained from dementia cohorts included in the Dementia Genetics Spanish Consortium (DEGESCO)^30^. At all sites AD diagnosis was established by a multidisciplinary working group, including neurologists, neuropsychologists and social workers, according to the DSM-IV criteria for dementia and the National Institute on Aging and Alzheimer’s Association’s (NIA– AA) 2011 guidelines for diagnosing AD. In the present study we considered as AD cases any individuals with dementia diagnosed with probable or possible AD at any point in their clinical course. For further details on the contribution of the sites, see Supplementary Table 1. Written informed consent was obtained from all participants. The Ethics and Scientific Committees have approved this research protocol (Acta 25/2016, Ethics Committee. H., Clinic I Provincial, Barcelona, Spain).

**Table 1.** Results for the AD loci selected for follow-up. *Note: FreqA1 is from GR@ACE discovery dataset. P-value for significance < 5 × 10-8. Effect allele: Allele 1.*

| Discovery meta-analysis |  |  |  |  |  |  |  |  | Follow-up datasets |  | Overall |  |
| --- | --- | --- | --- | --- | --- | --- | --- | --- | --- | --- | --- | --- |
| Chr | Closest gene | SNP | BP | Allele 1 | Allele 2 | FreqA1 | OR[CI95%] | P | OR[CI95%] | P | OR[CI95%] | P |
| Variants showing novel genome-wide significant association with AD |  |  |  |  |  |  |  |  |  |  |  |  |
| 2 | PRKD3/NDUFAF7 | rs876461 | 37515958 | A | G | 0.143 | 1.07[1.04-1.09] | 9.14E-07 | 1.08[1.04-1.13] | 3.07E-04 | 1.07[1.05-1.09] | 1.34E-09 |
| 8 | SHARPIN | rs34674752 | 145154222 | A | G | 0.052 | 1.11[1.06-1.16] | 4.02E-06 | 1.20[1.10-1.31] | 1.65E-05 | 1.13[1.09-1.18] | 1.00E-09 |
| 8 | SHARPIN | rs34173062 | 145158607 | A | G | 0.085 | 1.16[1.11-1.21] | 1.33E-11 | 1.09[1.02-1.17] | 7.35E-03 | 1.14[1.10-1.18] | 9.62E-13 |
| 16 | PLCG2 | rs3935877 | 81900853 | C | T | 0.868 | 0.92[0.90-0.95] | 1.12E-07 | 0.92[0.85-0.99] | 1.96E-02 | 0.92[0.90-0.95] | 6.85E-09 |
| 17 | CHRNE | rs72835061 | 4805437 | A | C | 0.085 | 1.09[1.06-1.12] | 3.92E-09 | 1.07[1.02-1.12] | 7.83E-03 | 1.09[1.06-1.11] | 1.51E-10 |
| 21 | APP | rs2154481 | 27473875 | C | T | 0.483 | 0.95[0.93-0.96] | 9.26E-10 | 0.96[0.93-0.99] | 3.31E-03 | 0.95[0.94-0.96] | 1.39E-11 |
| Previously reported genome-wide significant hits replicating in the follow-up |  |  |  |  |  |  |  |  |  |  |  |  |
| 4 | HS3ST1 | rs4351014 | 11027619 | C | T | 0.684 | 0.94[0.92-0.96] | 5.37E-10 | 0.93[0.88-0.98] | 4.54E-03 | 0.94[0.92-0.95] | 9.16E-12 |
| 16 | IL34 | rs4985556 | 70694000 | A | C | 0.111 | 1.08[1.05-1.11] | 2.28E-08 | 1.09[1.03-1.16] | 4.59E-03 | 1.08[1.06-1.11] | 3.91E-10 |
| 16 | PLCG2 | rs12444183 | 81773209 | A | G | 0.407 | 0.95[0.93-0.97] | 1.48E-08 | 0.92[0.88-0.96] | 3.23E-05 | 0.95[0.93-0.96] | 6.81E-12 |
| Suggestive signals (not replicating) |  |  |  |  |  |  |  |  |  |  |  |  |
| 14 | ELK2AP | rs7153315 | 106195719 | C | G | 0.750 | 0.94[0.92-0.96] | 9.80E-08 | 1.16[1.01-1.33] | 0.0412 | 0.94[0.92-0.97] | 9.04E-07 |
| 15 | SPPL2A | rs76523702 | 51002342 | C | T | 0.802 | 1.06[1.04-1.08] | 6.86E-08 | 1.02[0.97-1.07] | 0.3501 | 1.05[1.03-1.08] | 1.08E-07 |

#### Genotyping, quality control and imputation

DNA was extracted from peripheral blood. Genotyping was conducted using the Axiom 815K Spanish Biobank array (Thermo Fisher) at the Spanish National Centre for Genotyping (CeGEN, Santiago de Compostela, Spain) (Supplementary information, methods). We conducted previously described standard quality control prior to imputation^28^. In brief, individual quality control includes genotype call rates > 97%, sex checks and no excess heterozygosity; we removed population outliers as well (European cluster of 1000 Genomes). We included variants with a call rate > 95%, with minor allele frequency (MAF) > 0.01, in Hardy-Weinberg equilibrium (p < 1×10^−4^ in controls) and without differential missingness between cases and controls (Supplementary Table 2, Supplementary Figure 1). Imputation was carried out using the Haplotype reference consortium^31^ (HRC, full panel) and the 1000 Genomes reference panel^32^ (for indels only) on the Michigan Imputation server (https://imputationserver.sph.umich.edu). Rare variants (MAF < 0.001) and variants with low imputation quality (*R*^*2*^ *<* 0.30) were excluded. Logistic regression models, adjusted for the first four ancestry principal components^28^, were fitted using Plink (v2.00a). Population-based controls were used; therefore, age was not included as a covariate (Supplementary Table 3). After quality control steps, we included 6,331 AD cases and 6,055 control individuals and tested 14,542,816 genetic variants for association with AD.

**Table 2.**
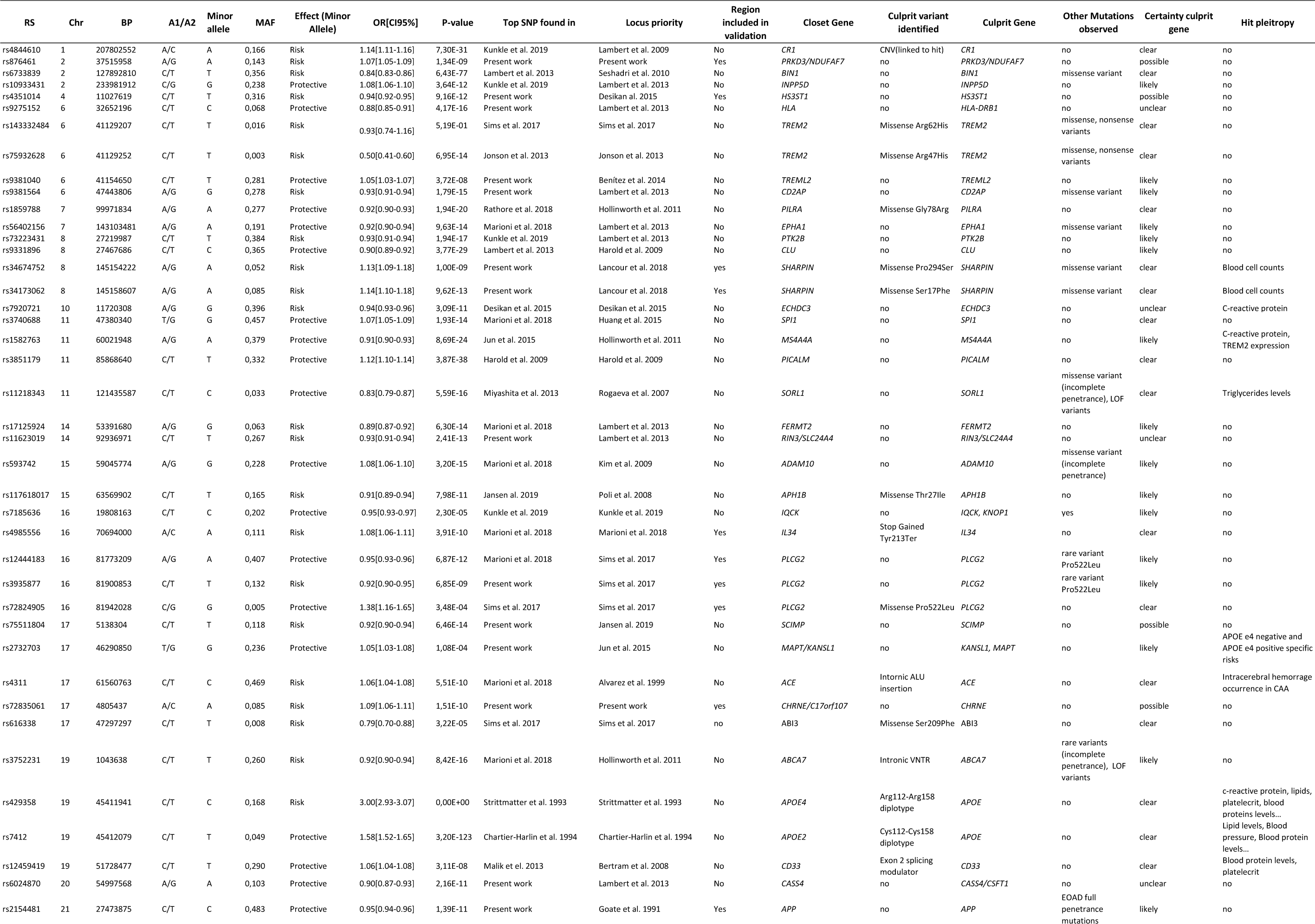
The genetic landscape of late-onset Alzheimer’s Disease. *Note: Association results of overall meta-GWAS cohorts. OR: effect allele A1. Allelic Freq. is from A1 in GR@ACE discovery dataset. LOF: Loss-of-function.*

#### IGAP summary statistics

GWAS summary results from the IGAP were downloaded from the National Institute on Aging Genetics of Alzheimer’s Disease Data Storage Site (NIAGADS, https://www.niagads.org/)^29^. Details on data generation and the analyses by IGAP have been previously described^29^. In brief, IGAP is a large study based upon genome-wide association using individuals of European ancestry. Stage 1 of IGAP comprises 21,982 Alzheimer’s disease cases and 41,944 cognitively normal controls from four consortia: the Alzheimer Disease Genetics Consortium (ADGC), the European Alzheimer’s Disease Initiative (EADI), the Cohorts for Heart and Aging Research in Genomic Epidemiology Consortium (CHARGE) and the Genetic and Environmental Risk in AD Consortium Genetic and Environmental Risk in AD/Defining Genetic, Polygenic and Environmental Risk for Alzheimer’s Disease Consortium (GERAD/PERADES). Summary statistics are available for 11,480,632 variants, both genotyped and imputed (1000 Genomes phase1v3). In Stage 2, 11,632 SNPs were genotyped in an independent set of 8,362 Alzheimer’s disease cases and 10,483 controls.

#### UK Biobank summary statistics

UK Biobank data, including health, cognitive and genetic data, were collected on over 500,000 individuals aged 37–73 years from across Great Britain (England, Wales and Scotland) at the study baseline (2006–2010) (http://www.ukbiobank.ac.uk)^33^. Several groups have demonstrated the utility of self-report of parental history of AD for case ascertainment in GWAS (Proxy–AD approach)^7,9,10^. For this study we used the published summary statistics of Marioni et al.^9^. They included, after stringent quality control, 314,278 unrelated individuals for whom AD information was available on at least one parent in UK Biobank (https://datashare.is.ed.ac.uk/handle/10283/3364). In brief, the genetic data of 27,696 participants whose mother had dementia (maternal cases) were compared with the 260,980 participants whose mother did not have dementia. Likewise, the 14,338 participants whose father had dementia (paternal cases) were compared with the 245,941 participants whose father did not have dementia^9^. The phenotype of the parents is independent, and therefore the estimates could be meta-analysed. After analysis, the effect estimates were made comparable to a case-control setting. Further information on the transformation of the effect sizes can be found elsewhere^9,34^. The data available comprises summary statistics of 7,794,553 SNPs imputed to the HRC reference panel (full panel).

### Data analysis

After study-specific variant filtering and quality-control procedures, we performed a fixed-effects inverse-variance-weighted meta-analysis^35^ on the summary statistics mentioned above.

To determine the variants with the strongest association per genomic region, we performed clumping on SNPs with a genome-wide significant *p-*value (*p* < 5×10^−8^) (Plink v1.90, maximal linkage disequilibrium (LD) with R^2^ < 0.001 and physical distance 1 Mb). In the *APOE* region, the significance level (*p* ∼ 0 for multiple SNPs) interferes with clumping; therefore, only the variants determining the *APOE ε4* and *APOE ε2* alleles were kept in this region^36^ (rs429358 and rs7412). LD information was calculated using the GR@ACE imputed genotypes as a reference. Chromosomal regions associated with AD in previous studies were excluded from follow-up (Lambert et al.^12^, Kunkle et al.^6^, Jansen et al.^10^). We also performed a functional annotation using FUMA^37^ (see ‘Supplementary Methods’).

#### Confirmation of loci containing novel associations with AD loci

We searched for independent evidence of association with AD for the variants with suggestive association (*p* < 10^−5^) located in proximity (200 Kb) to nine loci selected for follow-up. This strategy was adopted in order to allow for potential refinement of the top associated variants during the replication effort. For this confirmatory experiment we studied 19,089 AD cases and 39,101 controls not used in the GRA@CE or IGAP studies from 16 additional cohorts many of them collected and analysed by the European Alzheimer Disease Biobank project (JPND-EADB). See Supplementary Table 3 for details of AD cases included in this study and their origin and Supplementary Information for descriptions of the replication cohorts). Logistic regression models were fitted with a minimum of four principal ancestry components to correct for the population substructure. Inverse variance weighted meta-analysis was performed on all datasets in both discovery and follow-up stages.

Conditional analyses were performed in regions where multiple variants were associated with AD using logistic regression models adjusting for the genetic variants in the region. In the chr17-*SCIMP* region we adjusted rs75511804 for rs72835017, and in the chr16-*PLCG2* region we adjusted rs12444183 for rs3935877 and rs72824905.

#### Polygenic Risk Scores

##### Validation of PRS in clinical and pathologically confirmed AD cases

We calculated a weighted individual PRS based on the 39 genetic variants that showed genome-wide significant evidence of association with AD in the present study (Figure 2, Supplementary Table 4). Selected variants were directly genotyped or imputed with high quality (median imputation score R^2^ = 0.93). PRS were generated by multiplying the genotype dosage of each risk allele for each variant by its respective weight, and then summing across all variants. We weighted by the effect size from previous IGAP studies (Kunkle^6^ (36 variants), Sims^16^ (2 variants), Jun^38^ (*MAPT* locus), Supplementary Table 4).

**Figure 2:**
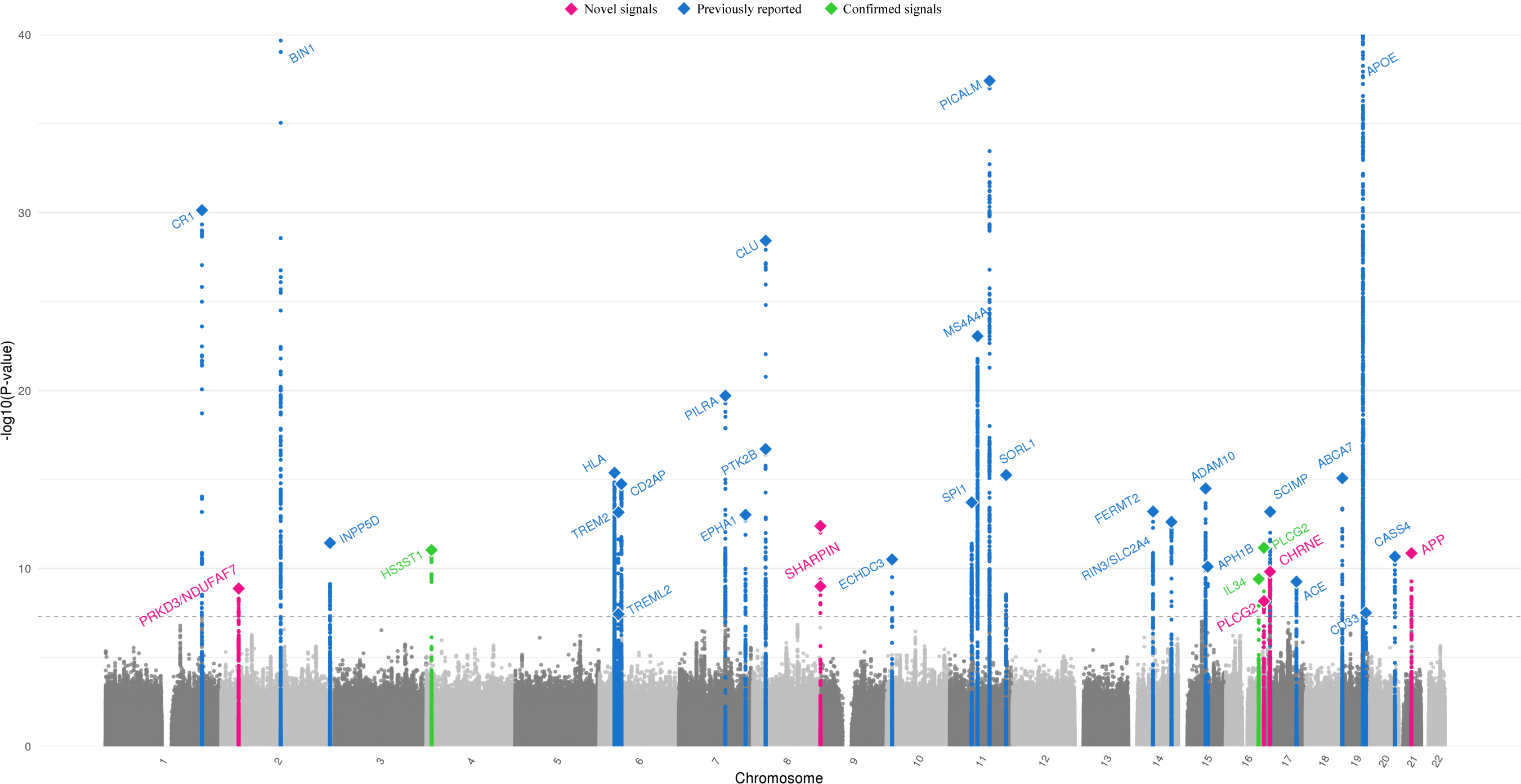
Manhattan plot of overall meta-analysis for genome-wide association in Alzheimer’s disease.

We validate the PRS in a sample of 676 AD cases diagnosed based on clinical criteria and 332 pathologically confirmed AD cases from the European Alzheimer’s Disease Biobank (EADB)–Fundació ACE/Barcelona Brain Bank dataset (EADB–F.ACE/BBB, Supplementary information). This dataset was not used in any genetic study. In this dataset all pathologically confirmed cases were scored for the presence or absence of concomitant pathologies. In all analyses we compared the AD patients to the same population-based control dataset (n = 1,386). We performed analyses to test the robustness of the PRS. We first tested the effect of adding additional variants below the genome-wide significance threshold using a pruning and thresholding approach. For this we used the summary statistics of IGAP^29^ study and we selected independent variants using the *clump_data()* function from the TwoSampleMR package (version 0.4.25). We used standard settings for clumping (R^2^=0.001 and window=1MB) and increasing p-value thresholds (>1×10^−7^, >1×10^−6^, >1×10^−5^, >1×10^−4^, >1×10^−3^, >1×10^−2^). We tested the association of the resulting with clinically diagnosed AD patients and pathologically confirmed AD patients. To evaluate the effect of diagnostic certainty we tested if the PRS was different between these two AD groups. For the PRS with 39 genome-wide significant variants we tested if there were sex-specific effects of the PRS, if the effect was different age-of-onset groups of AD and the effect of the PRS in the presence of concomitant brain pathologies.

##### Risk stratification of the validated PRSs

After validation of the PRS we searched for the groups at largest risk of AD in the large GR@ACE dataset (6,331 AD cases and 6,055 controls). We stratified the population into PRS (percent)tiles taking into account survival bias that is anticipated at old age^27^. To eliminate selection bias calculated the boundaries of the percentiles in the control participants aged 55 years and younger (n = 3,546). Based on the boundaries from this population the rest of the controls and all AD cases were then assigned into their appropriate percentiles. We first explored risk stratification using only the PRSs. For this we split the PRSs into 50 groups (2-percentiles) and compared all groups of subjects with the group that had the lowest PRS. Secondly, we explored risk stratification considering both *APOE* genotypes and the PRSs. The *APOE* genotypes were pooled in analyses as *APOE ε22/ε23* (n = 998, split into 7 PRS groups), *APOE ε33* (n = 7,611, split into 25 PRS groups), *APOE ε24/ε34* (n = 3,399, split into 15 PRS groups) and *APOE ε44* (n = 382, split into 3 PRS groups). We studied the effect of PRS across groups of individuals stratified by *APOE* genotypes with the lowest PRS group (*APOE* as the reference group using logistic regression models adjusted for four population ancestry components). Finally, we compared the median age at onset using a Wilcoxon test. All analyses were done in R (version 3.4.2).

## Results

### Genome-wide association study

We performed a meta-analysis of the summary statistics of the GR@ACE study (6,331 AD cases and 6,055 controls), the IGAP consortium (up to 30,344 AD cases and 52,427 controls) and the UK Biobank AD-proxy study (27,696 cases of maternal AD with 260,980 controls and 14,338 cases of paternal AD with 245,941 controls (Figure 1, Supplementary Table 3). Although we observed inflation in the resulting summary statistics (λ median = 1.08; see Supplementary Figure 2), it was not driven by an unmodelled population structure (LD score regression intercept = 1.036). We compared the results obtained to a second meta-analysis using only the case-control datasets (IGAP Stage 1–2 and GR@ACE datasets as a sensitivity analysis to identify false negative results due to possible dilution by the by-proxy approach in the UK Biobank (Supplementary Table 5). We identified a genome-wide significant association (*p* < 5×10^−8^) for 36 independent genetic variants in 35 genomic regions and two additional suggestive associations. The *APOE* locus contained two independent signals corresponding to the ε4 and ε2 alleles, respectively. The meta-analysis, including the by-proxy summary statistics, identified 11 additional loci reaching genome-wide significance with respect to case-control-only results. The incorporation of by-proxy summary statistics did not show an association in two previously reported AD loci (rs7185636-*IQCK* and rs386572859-*MAPT*) by the IGAP consortium^29,38^ and replicated in the GR@ACE dataset (OR = 0.93 [0.90-0.95], *p* = 4.5×10^−8^ and OR = 0.81 [0.75-0.87], *p*=7.9×10^−9^, respectively). We observed high correlation between the effect estimates from the case-control and by-proxy approach for the significant loci (R^2^ = 0.994, *p* = 8.1×10^−37^; Supplementary Figure 3).

**Figure 3:**
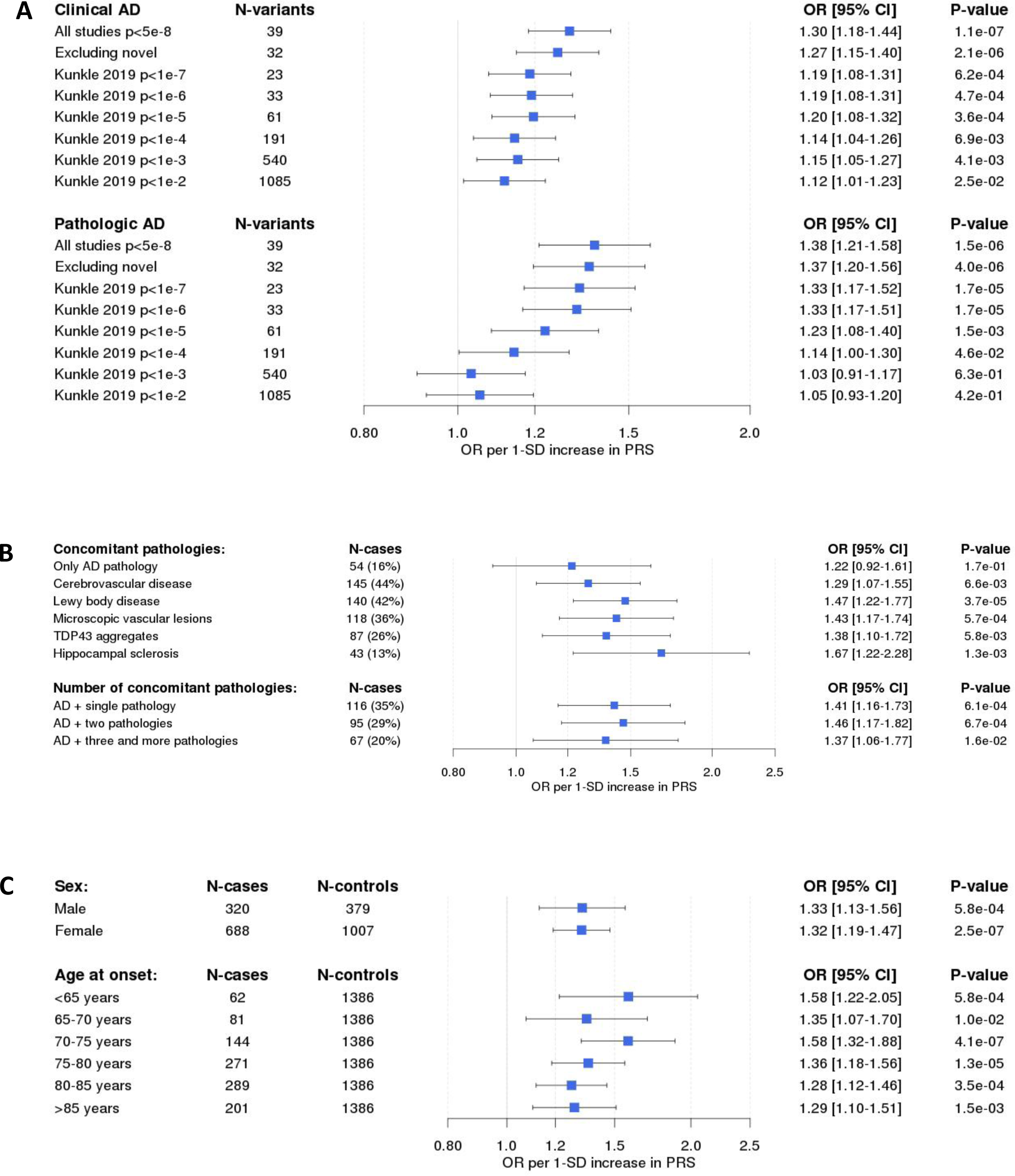
Clinical impact of a Polygenic Risk Score for Alzheimer’s Disease.

Among the 36 detected genome-wide significant (GWS) variants, 31 variants (86%) were reported in previously published studies or were in complete LD with known AD loci (Supplementary Table 5). We followed up the four novel genetic regions. We also followed up five genomic regions of interest, which were still awaiting replication (*PLCG2* and *IL-34*)^9^, in incomplete linkage disequilibrium with top SNPs previously reported loci^10,39^ (a locus near the *HS3ST1*^20^ gene) or showed only suggestive level of association (*p* < 1×10^−7^; *ELK2AP* and *SPPL2A*). We tested all variants in these nine genomic loci reaching suggestive level of association (*p <* 1×10^−5^) in the replication cohorts using 16 independent European-based cohorts (n cases = 19,087 and n controls = 39,101). In these nine genomic regions we tested 384 variants in the follow-up (Supplementary Table 6).

In the follow-up data found association signal in the same direction as the discovery in seven out of the nine genomic regions (Table 1, Supplementary Table 6 and Supplementary Figures 4–12). We combined discovery and follow-up and found in these seven genomic regions, nine independent genetic variants that reached GWS significance for association with AD (Table 1). Four of the genomic regions were not previously associated with AD and those are marked in the Manhattan-plot (Figure 2). We briefly discuss the results below.

**Figure 4:**
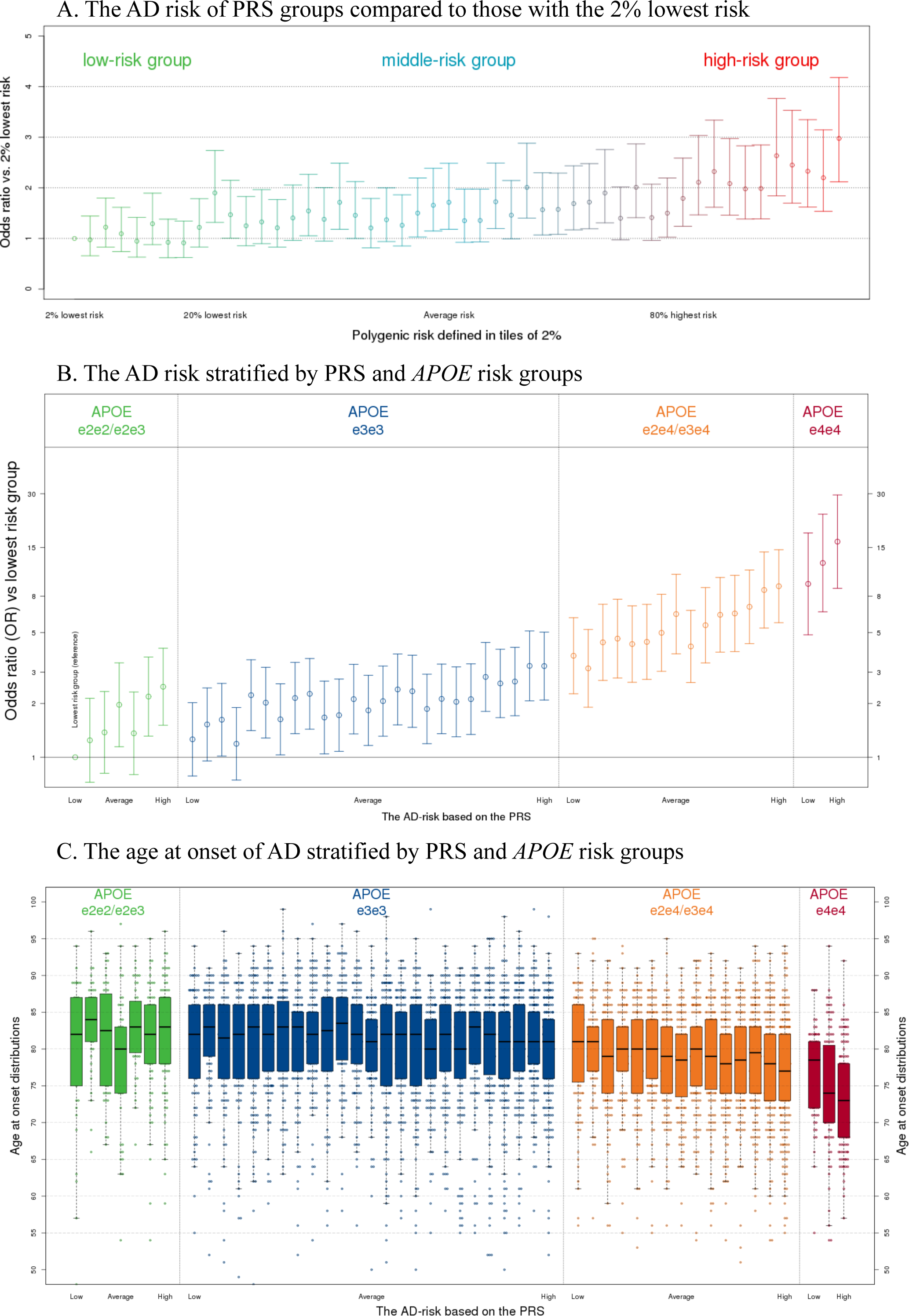
Risk of Alzheimer’s disease stratified by Polygenic Risk Score, *APOE* genotypes, and age at onset stratified by both.

Two genomic regions contain strong candidate AD genes (*APP* and *SHARPIN*) but variants in the region had not reached genome-wide significance in earlier meta-GWAS^15,35^. In *APP* we identified a very common (MAF = 0.46) intronic variant that associated with a reduced risk of AD (rs2154481, OR = 0.95 [0.93–0.96], *p* = 9.3×10^−10^). In *SHARPIN* we found two missense mutations (p.Ser17Phe and p.Pro294Ser) that are in linkage equilibrium (R^2^ = 1.3×10^−6^, D’ = 0.014 and *p* = 0.96). The *SHARPIN* p.Ser17Phe (MAF = 0.085) was significant in the discovery as well as the follow-up. In all data combined we found the variant associated with a 1.14-fold increased risk of AD (rs34173062, 95%CI 1.10–1.18, *p* = 9.6×10^−13^). *SHARPIN* p.Pro294Ser (MAF = 0.052) was significant after meta-analysing all data (rs34674752, OR = 1.13 [1.09–1.18], *p* = 1.0×10^−9^). The fourth variant is in a genomic region at chromosome 2, close to the genes *PRKD3* and *NDUFAF7*. The variant rs876461 (MAF = 0.143) was borderline significant in discovery (OR = 1.07 [1.04-1.09], *p* = 9.1×10^−7^) but emerged as the most significant variant in the region after in the combined analysis (OR = 1.07 [1.05-1.09], *p* = 1.3×10^−9^). The fifth association was in the 3’-UTR region of *CHRNE* (Cholinergic Receptor Nicotinic Epsilon Subunit). rs72835061 (MAF = 0.085) associated with a 1.09-fold increased risk of AD (95%CI 1.06-1.11, *p* = 1.5×10^−10^). This variant is relatively close to the *SCIMP*^10^ locus (333 Kb) and was in weak LD (R^2^ = 0.139, D’ = 0.446 and *p* < 0.0001) with the *SCIMP* top variant (rs75511804-*SCIMP*). A conditional analysis, including GR@ACE and the independent follow-up (Supplementary Table 7), showed similar effect sizes for rs72835017 after adjustment for rs75511804-*SCIMP* (and vice versa), in line with two independent signals that are in weak LD.

We strengthened the evidence of association with AD for three genomic regions. First, rs4351014 with AD (combined-OR = 0.94 [0.92-0.95], *p* = 9.2×10^−12^). This variant is in a gene poor region but has previously been linked to *HS3ST1*. A stop codon mutation (rs4985556, Tyr213Ter, MAF = 0.111) in the interleukin 34 (*IL34*) gene was previously reported to be associated in a by-proxy approach^9^. We confirmed this associated with an increased AD risk in both discovery and follow-up (combined OR = 1.08 [1.06-1.11], *p* = 3.9×10^−10^). The genomic region that contains the *PLCG2* gene has been associated with AD twice (the rare missense variant p.P522R in the *PLCG2* gene^16^ and rs12444183 near the promotor region of *PLCG2*^9^). After combination of discovery and follow-up a third independent association signal emerged in the *PLCG2* region (rs3935877, effect allele frequency = 0.868, OR = 0.92 [0.90-0.95], *p* = 6.9×10^−9^). We also strengthen the association of *PLCG2*-rs12444183 with AD (MAF = 0.407, combined-OR = 0.95[0.93-0.96], *p* = 6.8×10^−12^). Conditional analyses in the *PLCG2* region showed the association signal of all three variants (including the missense variant p.P522R in *PLCG2*^16^) in the *PLCG2* locus are independent (Supplementary Table 8).

The two loci reaching suggestive evidence of association (Table 1) in the discovery (rs7153315-*ELK2AP* and rs76523702-*SPPL2A*) did not replicate in the follow-up (rs76523702-*SPPL2A, p* = 0.35) or showed association in the opposite direction (rs7153315-*ELK2AP*, OR_discovery_ = 0.94 *vs* OR_follow-up_ = 1.16, Table 1).

To link the novel variants to specific genes and functional motifs in their genomic regions, we applied different strategies implemented on the FUMA platform (see ‘Supplementary Methods’). The selected candidate genes were implicated at least in three mapping strategies using this approach (*APP, IL34, CHRNE, PLCG2* and *SHARPIN*, Supplementary Tables 9–12, Supplementary Figure 13).

#### Validation of PRS for dementia

In total, after combined meta-analysis, 39 variants have been associated with AD at genome-wide significance (excluding the *APOE* region). We used these 39 variants and IGAP weighted effects to construct a PRS (Supplementary Table 4).

The PRS showed a stronger association with the pathologically confirmed AD cases (OR = 1.38, per 1-SD increase in the PRS, 95% CI [1.21–1.58]) than with clinical AD cases (OR = 1.30, 95%CI [1.18–1.44]) (Figure 3A). However, this difference was not statistically significant. We then investigated whether adding additional variants below the genome-wide significance threshold would lead to increased performance of the PRS. In both pathological and clinical AD cases, the association (as measured by the *p*-value of the effect estimate from logistic regression models) of the PRS decreased when adding to PRS variants below the conventional GWAS significance threshold (Figure 3A).

Concomitant brain pathologies were present in 84% of histopathological confirmed cases, and the PRS was associated AD cases with all tested concomitant pathologies. The strongest risk increase per 1-SD of the PRS was observed with concomitant hippocampal sclerosis (OR = 1.67, 95%CI [1.22–2.28]), Figure 3B). The smallest effect for the PRS was observed in the 16% of cases that had only AD pathology (OR = 1.22, 95%CI [0.92– 1.61]). The AD patients in our series often had more than one concomitant pathology (48.8%), but there was no difference in the effect estimate of the PRS when more than one pathology was present (Figure 3B).

Finally, we investigated the differential effect of sex and age at onset on the effect estimate of the PRS (Figure 3C). The effect of the PRS was comparable in males (OR = 1.33 per 1-SD, *p =* 5.8×10^−4^) and females (OR = 1.32 per 1-SD, *p =* 2.5×10^−7^). Overall, there were significant effects of the PRS on AD risk in all five-year age-at-onset groups. The strongest effect was observed in the group with an age at onset of 70–75 (OR = 1.58, per 1 SD, *p =* 4.1×10^−7^).

#### Risk stratification using polygenic risk scores and APOE

In the independent AD cohort, the PRS of 39 variants showed the strongest association with AD (Figure 3). We therefore used this PRS in the large GR@ACE dataset (6,331 AD cases and 6,055 controls) to identify those at highest genetic risk of AD. Overall, the PRS was associated with a 1.27-fold (95% CI 1.23–1.32) increased risk for every standard deviation increase in the PRS (*p* = 7.3×10^−39^). When we stratified all individuals into 2% percentiles of the PRS, we observed a gradual risk increase over the 2% percentile groups (Figure 4A, Supplementary Table 13), and when comparing the extremes, those with the 2% highest risk had a 2.98-fold (95% CI 2.12–4.18) increased risk compared to those with the 2% lowest risk (*p* = 3.2×10^−10^).

We then studied the ability of the PRS to identify high individuals at risk of the subjects within and across *APOE* genotype groups. The *APOE* categories were split into PRS subgroups depending on number or subjects available: seven PRS groups for the *APOE ε22/ε23* genotype, 25 for *APOE ε33*, 15 for *APOE ε24/ε34* and three for *APOE ε44*. Within each *APOE* genotype category we found that the group of individuals with the highest PRS score had a higher risk compared to the lowest scored group (Figure 4B).

There was a 2.48-fold increased risk for *APOE ε22/ε23* (*p* = 3.4×10^−4^), 2.67-fold for *APOE ε33* (*p* = 3.5×10^−9^), 2.47-fold for *APOE ε24/ε34* (*p* = 6.8×10^−6^) and 2.02-fold for *APOE ε44* (*p* = 3.4×10^−2^). The PRS is able to modify the risk associated with *APOE* such that *APOE ε22/ε23* carriers with the highest PRS has a significantly higher risk than *APOE ε33* carriers in the lowest scored group (*p* = 7.8×10^−4^), *APOE ε33* carriers in the highest PRS group has a risk comparable to *APOE ε4* heterozygote carriers with the lowest PRS (*p* = 0.40), and *APOE ε4* heterozygotes with the highest PRS was not significantly different from *APOE ε4* homozygotes with the lowest PRS (*p* = 0.68). Finally, we compared the risk extremes and found a 16.2-fold (95% CI 8.84–29.5, *p* = 1.5×10^−19^) increased risk for the highest-PRS group compared to the lowest-PRS group (the highest PRS *APOE ε44* group vs the lowest PRS *APOE ε22/ε23* group) (Supplementary Table 14).

The stratification of individuals by PRS and *APOE* genotype also influences age at onset. We found a significant (*p*_*Wilcoxon*_ = 1.7×10^−6^) difference of 9 years in the median age at onset in individuals with the lowest PRS (the median onset is 82 years for *APOE ε22/ε23* with the lowest PRS) compared with individuals with the highest risk (the median onset is 73 years for *APOE ε44* with the highest PRS risk) (Figure 4C). The PRS did not determine age at onset in either *APOE ε22/ε23* (lowest = 82 years, highest = 83 years, *p*_*Wilcoxon*_ = 0.39) or *APOE ε33* (lowest = 82 years, highest = 81 years, *p* = 0.16). By contrast, in the *APOE ε4* heterozygotes, the PRS determined a 4-year difference (*p*_*Wilcoxon*_ = 6.9×10^−5^) in median age at onset between the lowest-risk (81 years) and highest-risk (77 years) tiles. Moreover, although the PRS did not have a significant effect on risk within *APOE ε4* homozygotes, it did determine a 5.5-year difference in median age at onset (*p*_*Wilcoxon*_= 4.6×10^−5^) between the low-risk (median = 78.5 years) and high-risk (73 years) tiles.

## Discussion

In the present work we report on the largest meta-GWAS for AD risk to date comprising genetic information of 467,623 individuals of European ancestry. Using our meta-GWAS approach and follow-up analysis, we identified four genomic regions that are significantly associated with the risk of AD for the first time (in *CHRNE, APP, SHARPIN* and near *PRKD3/NDUFAF7*). We also strengthen the association of the *PLCG2* region with AD by the identification of an additional association signal in the region. This brings the total of genetic variants associated with AD to 39 (excluding *APOE* variants and the very rare coding variants). We validated a PRS based on these variants and show that this PRS can identify individuals at highest genetic risk of AD independent of *APOE*. In fact, with this PRS we can identify *APOE ε3* homozygote carriers who have comparable risk for AD as *APOE ε4* heterozygote carriers, as well as *APOE ε4* heterozygote carriers who have comparable risk for AD as *APOE ε4* homozygotes carriers. We also demonstrate that the PRS determines a 4 to 5.5-year age-at-onset difference within *APOE ε4* carriers. We conclude that the PRS of the currently known genetic risk of AD captures important differences in AD risk and age at onset. The effects of both risk stratification and age at onset are important for research studies and clinical trials.

The work presented builds on a long, ongoing global effort of genetic researchers to identify genetic loci associated with AD using family-based studies and genome-wide association studies. We have summarised the landscape of AD genetics of the last 30 years in Figure 5. We can observe that with the increasing sample sizes across studies, more and more variants are found. After review of the literature, we estimate that the culprit variant is known in only 15 (40%) out of 39 loci (Table 2). Therefore, much is to be gained from candidate gene studies in these other loci. With the current work we have added six novel genetic variants in five genetic loci to the landscape; they are discussed below.

**Figure 5.**
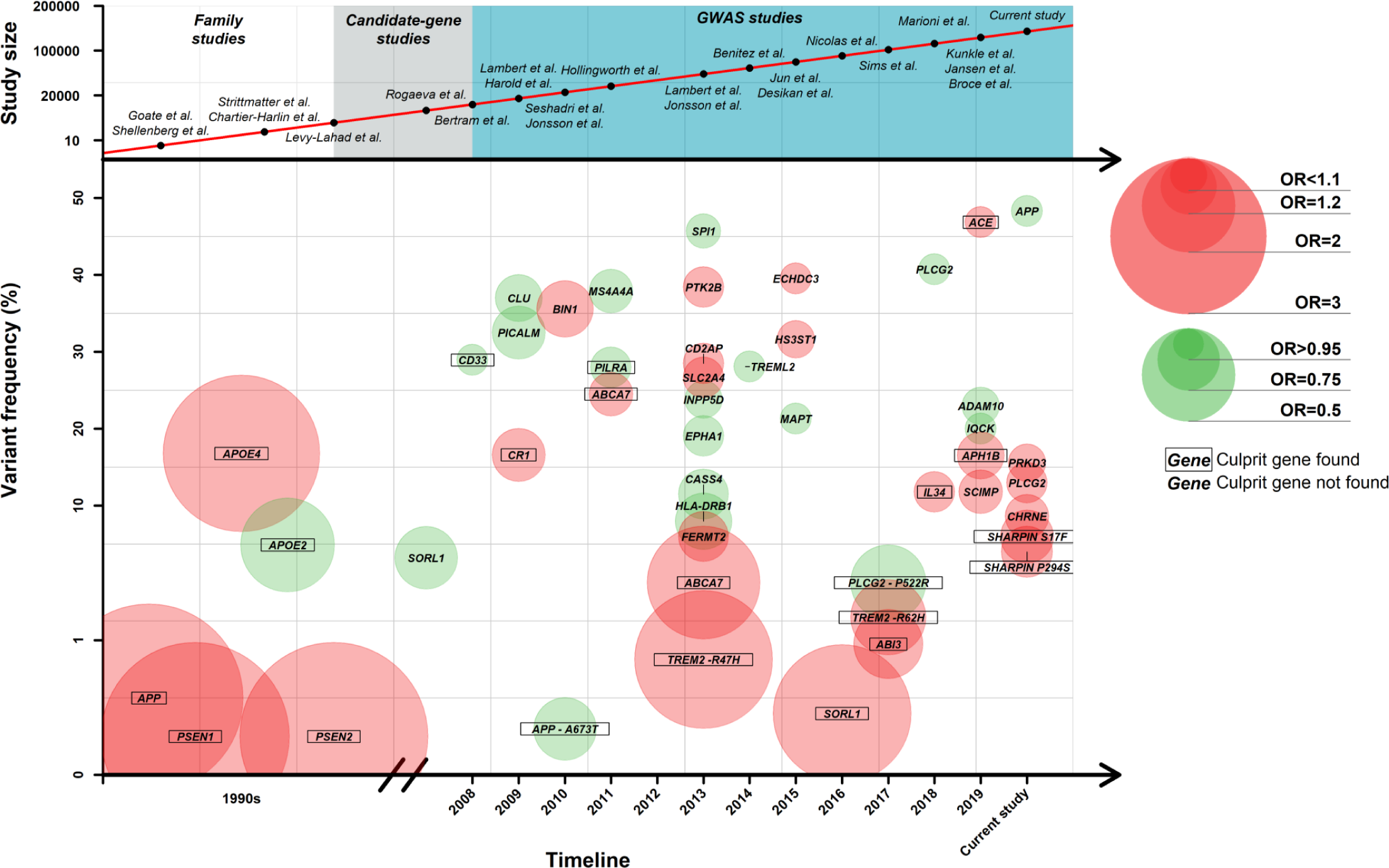
Landscape for Alzheimer’s Disease over the last years. GWAS= Genome-Wide Association Study; OR=Odds Ratio

The most interesting finding of this work was the confirmation of a very common variant in the *APP* locus (rs2154481, MAF (C-allele) = 0.483). The C-allele of this SNP confers subtle protection against AD to the carriers (OR = 0.95 [0.94-0-96], *p* = 1.39×10^−11^). The genetic marker is in a DNase hypersensitive area of 295 bp (chr21:27473781-27474075) probably involved in the transcriptional regulation of the *APP* gene. Indeed, the variant is an eQTL for the *APP* mRNA and antisense transcript of the *APP* gene named AP001439.2 in public eQTL databases^40^ (Supplementary Figure 14). Importantly in the dorsolateral prefrontal cortex of 726 individuals^41^, the protective variant increased expression of the *APP* transcript.

Additional functional evidence supporting the role of this region in the modulation of the *APP* transcript has been published recently^42^. Specifically, Craig et al. describe a block of 13 SNPs within the *APP* locus associated with intellectual abilities in children. This LD block, including our top hit for AD (rs2154481), was associated with intelligence (IQ) in children from the Avon Longitudinal Study of Parents and Children (ALSPAC, n = 5,165, beta = 1.36, *p* = 3.5×10^−5^ for the rs2154481 C-allele). The authors suggest that this LD block within *APP* is involved in the control of gene expression in this locus. Using EMSA and luciferase reporter assays, they demonstrate that the C-allele of a linked SNP (rs2830077) increased the TFCP2 transcription factor avidity to its binding site and increased the enhancer activity of this specific intronic region^42^. Interestingly, the C-allele of the proposed marker (rs2830077) is in high LD with the C-allele of rs2154481 detected in our study (LDlink, D’= 0.94, R^2^ = 0.87, p < 0.0001). The results support increased *APP* gene expression. It is not necessary to mention that the *APP* locus has been one of the major drivers of the classic amyloid hypothesis for AD^43,44^. Now, our results reconcile autosomal dominant and complex Alzheimer disease genetic causality. The incontrovertible demonstration of the existence of the common variant within the *APP* associated with sporadic AD and full penetrance mutations affecting autosomal dominant AD strongly supports a common causal path. Hence, our results ultimately confirm the role of *APP* physiology in not only early-onset Alzheimer’s disease but also late-onset AD, as recently proposed by the IGAP consortium^29^. Nevertheless, the expression results linked to the rs2154481 might appear somehow counterintuitive because increased expression of the *APP* mRNA appeared to be related to disease protection. This could be due to potential *hormesis* or U-effect properties of this locus, where discrete increases of the wild-type (non-mutated) *APP* transcript could be protective and increased expression of the mutant gene might be harmful^45^. The *hormesis* theory, if true, might help to explain the accelerated cognitive deterioration observed in AD patients treated to reduce beta-amyloid in their brain using beta-secretase inhibitors^46,47^. An alternative hypothesis proposes that rs2154481 mechanisms could be related to the overexpression of protective fragments of the APP protein^48^. Regardless, we feel that more basic research is needed to understand the observed association. We are confident that disentangling first the culprit variant and then the molecular mechanism associated with rs2154481 hit will help to refine the amyloid hypothesis.

We found two missense variants in *SHARPIN* associated with AD. Both variants have a relatively large effect (Figure 5), which strongly suggests that *SHARPIN* is the culprit gene in the locus. *SHARPIN* was proposed as a candidate gene for AD by prioritisation of GWAS signals using protein network analysis^49^ and by genetic association in the Japanese population^50^. In the Japanese study, a rare nonsynonymous variant, rs572750141 (NM_030974.3:p.Gly186Arg) was associated at suggestive significance with an increased risk of AD. The amino-acid change resulted in aberrant cellular localisation of the variant protein and attenuated the activation of NF-κB, a central mediator of inflammatory and immune responses. Furthermore, spontaneous mutations in the mouse *SHARPIN* gene resulted in immune system dysregulation^51^, and rs34173062 was associated with changes in blood cell indices—specifically myeloid white cell and compound white cell^52^ changes. Our findings confirm *SHARPIN* as an important AD gene, and the effect of the missense mutations on immunity can be directly studied in cell models.

We also found a *PRKD3/NDUFAF7* signal in our meta-GWAS, but the top hit shifted during the overall meta-analysis. Still, the locus retained genome-wide significance, requiring future independent replication. It is important to emphasise that it is an excellent candidate for further follow-up. The protein kinase D (PKD) family of serine/threonine protein kinases occupy a unique position in signalling pathways initiated by diacylglycerol and protein kinase C. PKDs are involved in resistance to oxidative stress, cell survival, migration, differentiation and proliferation^53^.

We confirmed an intronic variant in *CHRNE* gene (rs72835061). Functional analysis of the discovery dataset already supported *CHRNE* as the most likely culprit gene. In fact, the rs72835061 variant is a strong eQTL of *CHRNE* in which the allele A increases the expression in the brain and many other tissues according to GTEx (top differential expression in the frontal cortex, *p* = 2.1×10^−13^) (Supplementary Figure 15). The *CHRNE* locus encodes the *Homo sapiens* cholinergic receptor (AChR), nicotinic epsilon, which is expressed in muscles and associated with congenital myasthenic syndrome (fast-channel type)^54^. Congenital myasthenic syndromes (CMSs) are a group of rare genetic disorders of the neurological junction that can result in structural or functional weakness. A change in the ε subunit leads to an increase or decrease in AChR protein signalling, which impairs cell-to-cell communication in the neuromuscular junction^55^. The detection of a potential hypermorph allele linked to AD risk and affecting cholinergic function could re-introduce this neurotransmitter pathway into the search for preventative strategies. Further functional studies are needed to consolidate this hypothesis.

For the *PLCG2* locus we obtained independent replication for rs12444183 and new genome-wide significance for rs3935877. The rare protective missense mutation in *PLCG2*^16^ (p.Pro522Arg) and now two other different haplotypes around *PLCG2* associated with AD reinforce the role of this genomic region in AD susceptibility. The dissection of the molecular mechanism behind these potential regulatory variants might help to elucidate the gain or loss of the function mechanism of the *PLCG2* gene. This information could be critical for drug-targeting purposes^56^.

So far not much attention has gone to a loss-of-function variant (p.Tyr213Ter) in interleukin *IL34*. In a previous AD-by-proxy study, Marioni et al.^9^ suggested *IL34*-rs4985556, but independent replication was pending. Interleukin-34 is a cytokine that promotes the differentiation and viability of monocytes and macrophages through the colony stimulating factor 1 receptor (CSF-1R)^57^. It promotes the release of proinflammatory chemokines and, thereby plays an important role in innate immunity and inflammatory processes^58^. Furthermore, microglia treated with IL-34 attenuated the oligomeric amyloid-β neurotoxicity in primary neuron-microglia co-cultures^59^. Intracerebroventricular administration of IL-34 ameliorated the impairment of associative learning and reduced oligomeric amyloid-β levels in an *APP/PS1* transgenic mouse model of AD. Therefore, this stop codon could increase the risk of AD by reducing the neuroprotective properties of *IL34* against the neurotoxicity of oligomeric amyloid-β or by modulating microglia reactivity in AD brains. The finding suggests that the generation of recombinant IL-34-based therapies could be a promising strategy for combating AD^60^.

Finally, in this study, we explored the potential utility of the PRS using the extended panel of common genetic variants detected. We demonstrated how PRS modifies the risk and onset of AD within *APOE* genotypes. The carriers of high-risk individuals into clinical trials is relevant as *APOE ε4*. The risk modification by the joint effect of common variants was most pronounced in *APOE ε4* carriers, in whom there was a difference of up to 4 years in onset age between the low- and high-risk tiles of the PRS in *APOE ε34* and a 4.5-year difference in *APOE ε44*.

The added value of PRSs of common variants with small effects in terms of improved discrimination between AD cases and controls was reported previously as marginal^22,26,61,62^. Our study and others recently published^24,26,27^ showed, however, that their effects are substantial for risk and age at onset. We show that the PRS could be instrumental in clinical diagnosis for AD, the effect of the PRS was similar in men and women, and there is risk differentiation at all ages of onset. This shows the wide applicability and robustness of PRS. We did not observe that adding a sub-threshold signal from GWAS improved the PRS, as had been previously suggested^62^. The identification of subgroups at high genetic risk of AD with an earlier disease onset in the general population has important implications for precision medicine. Pathological changes related to AD begin to develop up to decades before the earliest clinical symptoms^63^. Therefore, preventive interventions are increasingly introduced in the subgroup of individuals with a high genetic risk at a younger age^19,20^ reducing the duration of the trials and thus giving them the opportunity to access the most promising treatments for high-risk individuals. In this context, *APOE ε4* homozygotes carriers are considered high risk, but as shown here the subjects with the highest PRS values and carrying one copy of *APOE ε4* have a similar risk (Figure 4B). This is relevant as this group represent ∼1% of our control population, the same percentage as all *APOE ε4* homozygotes. However, further validation on the effect of proposed PRS in longitudinal series of healthy and mild cognitive impairment (MCI) subjects, in context of other biomarkers (e.g. imaging) are mandatory to determine the predictive utility of this paradigm. With continued identification of culprit variants and additional risk loci, we anticipate that the precision of PRS will be further enhanced^64^.

In sum, the current work reinforces the importance of increasing the sample size in future meta-analyses to identify novel genetic associations with AD and refine known loci to converge on the culprit variants. We described six novel associations with AD of common alleles in or near the genes: *APP, PLCG2, CHRNE, SHARPIN* and *PRKD3/NDUFAF7*. These signals reinforce that AD is complex disease in which amyloid processing and immune response play key roles. We add to the growing body of evidence that polygenic scores of all genetic loci to date, in combination with *APOE* genotypes, are robust tools for predicting risk and age at onset of AD. These properties make PRS promising in selecting individuals at risk in order to apply preventative strategies.

## Data Availability

Not Applicable

## Acknowledgments

We would like to thank patients and controls who participated in this project. The present work has been performed as part of the doctoral program of I. de Rojas at the Universitat de Barcelona (Barcelona, Spain). The Genome Research @ Fundació ACE project (GR@ACE) is supported by Grifols SA, Fundación bancaria ‘La Caixa’, Fundació ACE, and CIBERNED. A.R. and M.B. receive support from the European Union/EFPIA Innovative Medicines Initiative Joint undertaking ADAPTED and MOPEAD projects (grant numbers 115975 and 115985, respectively). M.B. and A.R. are also supported by national grants PI13/02434, PI16/01861, PI17/01474 and PI19/01240. Acción Estratégica en Salud is integrated into the Spanish National R + D + I Plan and funded by ISCIII (Instituto de Salud Carlos III)–Subdirección General de Evaluación and the Fondo Europeo de Desarrollo Regional (FEDER–’Una manera de hacer Europa’). Some control samples and data from patients included in this study were provided in part by the National DNA Bank Carlos III (www.bancoadn.org, University of Salamanca, Spain) and Hospital Universitario Virgen de Valme (Sevilla, Spain); they were processed following standard operating procedures with the appropriate approval of the Ethical and Scientific Committee.

Amsterdam dementia Cohort (ADC): Research of the Alzheimer center Amsterdam is part of the neurodegeneration research program of Amsterdam Neuroscience. The Alzheimer Center Amsterdam is supported by Stichting Alzheimer Nederland and Stichting VUmc fonds. The clinical database structure was developed with funding from Stichting Dioraphte. Genotyping of the Dutch case-control samples was performed in the context of EADB (European Alzheimer DNA biobank) funded by the JPco-fuND FP-829-029 (ZonMW projectnumber 733051061). 100-Plus study: We are grateful for the collaborative efforts of all participating centenarians and their family members and/or relations. This work was supported by Stichting Alzheimer Nederland (WE09.2014-03), Stichting Diorapthe, horstingstuit foundation, Memorabel (ZonMW projectnumber 733050814) and Stichting VUmc Fonds. Genotyping of the 100-Plus Study was performed in the context of EADB (European Alzheimer DNA biobank) funded by the JPco-fuND FP-829-029 (ZonMW projectnumber 733051061). Longitudinal Aging Study Amsterdam (LASA) is largely supported by a grant from the Netherlands Ministry of Health, Welfare and Sports, Directorate of Long-Term Care. The authors are grateful to all LASA participants, the fieldwork team and all researchers for their ongoing commitment to the study.

This work was supported by a grant (European Alzheimer DNA BioBank, EADB) from the EU Joint Programme – Neurodegenerative Disease Research (JPND) and also funded by Inserm, Institut Pasteur de Lille, the Lille Métropole Communauté Urbaine, the French government’s LABEX DISTALZ program (development of innovative strategies for a transdisciplinary approach to Alzheimer’s disease). Full acknowledgments for the studies that contributed data can be found in the Supplementary Note. We thank the numerous participants, researchers, and staff from many studies who collected and contributed to the data.

We thank the International Genomics of Alzheimer’s Project (IGAP) for providing summary results data for these analyses. The investigators within IGAP contributed to the design and implementation of IGAP and/or provided data but did not participate in analysis or writing of this report. IGAP was made possible by the generous participation of the control subjects, the patients, and their families. The i–Select chips was funded by the French National Foundation on

Alzheimer’s disease and related disorders. EADI was supported by the LABEX (laboratory of excellence program investment for the future) DISTALZ grant, Inserm, Institut Pasteur de Lille, Université de Lille 2 and the Lille University Hospital. GERAD was supported by the Medical Research Council (Grant n° 503480), Alzheimer’s Research UK (Grant n° 503176), the Wellcome Trust (Grant n° 082604/2/07/Z) and German Federal Ministry of Education and Research (BMBF): Competence Network Dementia (CND) grant n° 01GI0102, 01GI0711, 01GI0420. CHARGE was partly supported by the NIH/NIA grant R01 AG033193 and the NIA AG081220 and AGES contract N01–AG–12100, the NHLBI grant R01 HL105756, the Icelandic Heart Association, and the Erasmus Medical Center and Erasmus University. ADGC was supported by the NIH/NIA grants: U01 AG032984, U24 AG021886, U01 AG016976, and the Alzheimer’s Association grant ADGC–10–196728.

This research has been conducted using the UK Biobank public resource obtained through the University of Edinburg Data Share (https://datashare.is.ed.ac.uk/handle/10283/3364).

